# Serum Vitamin D levels are associated with increased COVID-19 severity and mortality independent of whole-body and visceral adiposity

**DOI:** 10.1101/2021.03.12.21253490

**Authors:** Pablo Esteban Vanegas-Cedillo, Omar Yaxmehen Bello-Chavolla, Natalia Ramírez-Pedraza, Bethsabel Rodríguez Encinas, Carolina Isabel Pérez Carrión, María Isabel Jasso-Ávila, Jorge Carlos Valladares-García, Diana Hernández-Juárez, Arsenio Vargas-Vázquez, Neftali Eduardo Antonio-Villa, Monica Chapa-Ibarguengoitia, Alfredo Ponce de Leon, José Sifuentes-Osornio, Carlos A. Aguilar-Salinas, Roopa Mehta

**Affiliations:** Unidad de Investigación de Enfermedades Metabólicas, Instituto Nacional de Ciencias Médicas y Nutrición Salvador Zubirán (INCMNSZ); Department of Endocrinology and Metabolism, INCMNSZ, Instituto Nacional de Ciencias Medicas y Nutricion Salvador Zubiran; Research Division, Instituto Nacional de Geriatría, Instituto Nacional de Ciencias Medicas y Nutricion Salvador Zubiran; MD/PhD (PECEM) program, Faculty of Medicine, National Autonomous University of Mexico, Instituto Nacional de Ciencias Medicas y Nutricion Salvador Zubiran; Department of Radiology, INCMNSZ, Instituto Nacional de Ciencias Medicas y Nutricion Salvador Zubiran; Department of Infectious Diseases, INCMNSZ, Instituto Nacional de Ciencias Medicas y Nutricion Salvador Zubiran; Internal Medicine Division, INCMNSZ, Instituto Nacional de Ciencias Medicas y Nutricion Salvador Zubiran; Instituto Tecnologico y de Estudios Superiores de Monterrey Tec Salud, Instituto Nacional de Ciencias Medicas y Nutricion Salvador Zubiran; Division of Nutrition, INCMNSZ, Instituto Nacional de Ciencias Medicas y Nutricion Salvador Zubiran

**Keywords:** Vitamin D, COVID-19, SARS-CoV-2, Adiposity, Severe disease

## Abstract

**INTRODUCTION:** Coronavirus disease (COVID-19) is a global pandemic. Vitamin D deficiency has been associated with susceptibility to infectious disease. In this study, the association between COVID-19 outcomes and vitamin D levels in patients attending a COVID-19 reference center in Mexico City are examined.

**METHODS:** Consecutive patients with confirmed COVID-19 were evaluated. All patients underwent clinical evaluation and follow-up, laboratory measurements and a thoracic computerized tomography, including the measurement of epicardial fat thickness. Low vitamin D was defined as levels <20ng/mL (<50nmol/L) and deficient Vitamin D as a level ≤12ng/mL (<30nmol/L)

**RESULTS:** Of the 551 patients included, low vitamin D levels were present in 45.6% and deficient levels in 10.9%. Deficient Vitamin D levels were associated with mortality (HR 2.11, 95%CI 1.24-3.58, p=0.006) but not with critical COVID-19, adjusted for age, sex, body-mass index and epicardial fat. Using model-based causal mediation analyses the increased risk of COVID-19 mortality conferred by low vitamin D levels was partly mediated by its effect on D-dimer and cardiac ultrasensitive troponins. Notably, increased risk of COVID-19 mortality conferred by low vitamin D levels was independent of BMI and epicardial fat.

**CONCLUSION:** Vitamin D deficiency (≤12ng/mL or <30nmol/L), is independently associated with COVID-19 mortality after adjustment for visceral fat (epicardial fat thickness). Low vitamin D may contribute to a pro-inflammatory and pro-thrombotic state, increasing the risk for adverse COVID-19 outcomes.

## INTRODUCTION

Coronavirus Disease (COVID-19), caused by the novel coronavirus severe acute respiratory syndrome coronavirus 2 (SARS-CoV-2), has caused significant burden in healthcare systems world-wide. In Mexico, COVID-19 has caused a large number of deaths, primarily due to the large prevalence of cardio-metabolic diseases linked to adverse outcomes and additionally due to the impact of socio-demographic factors which impact healthcare access and quality of care across Mexico(1–4). SARS-CoV-2 spreads primarily by close contact with respiratory droplets from infected individuals and contaminated surfaces(5,6). SARS-CoV-2 infects cells using the angiotensin converting enzyme-2 (ACE-2) receptor; infection can produce an interstitial pneumonia that may progress to acute respiratory distress syndrome (ARDS) and death(7,8). COVID-19 severity has been shown to be modified by the presence of cardio-metabolic comorbidities as well as inflammatory markers which may reflect vascular or respiratory deterioration(9–12).

Vitamin D is a steroid hormone involved in essential physiological roles including preserving bone integrity, immunomodulation by stimulating innate immunity and tempering adaptive immunity, infectious disease prevention and cardiovascular health(13,14). It also acts on the renin angiotensin aldosterone (RAAS) system, inhibiting the angiotensin converting enzyme (ACE(7)). Several factors are known to influence vitamin D levels; lower levels are associated with ethnicity, variation in sun exposure due to higher latitudes, season, time of day, clothing, sunscreen use and skin pigmentation, age, lower sun exposure, obesity and chronic illnesses(15). Low levels of vitamin D have been associated with increased susceptibility to infectious disease, particularly respiratory tract infections. Several studies have explored the relationship between COVID-19 and vitamin D levels(16,17); however, concerns regarding residual confounding and the lack of mechanistic interpretations for the association of low Vitamin D levels with adverse COVID-19 outcomes requires further studies. Overall, pooled evidence suggests that high Vitamin D levels have been associated with reduced risk of adverse COVID-19 outcomes, which may suggest a beneficial role for Vitamin D in COVID-19(18). Nevertheless, evidence from randomized controlled trials have not shown benefit from Vitamin D supplementation in COVID-19 or other infections due to the high heterogeneity across studies and its systematic use requires further evaluation(19,20).

The presence of obesity results in decreased bioavailability of vitamin D, which is probably related to sequestration into adipose tissue. Furthermore, higher visceral fat content has been shown to be related to a higher incidence of vitamin D deficiency(21,22). Obesity and ethnicity are important risk factors for severe disease and are also known to modulate vitamin D levels. This may be particularly relevant in Mexico, where high rates of diabetes and obesity have been associated with an increased risk of severe COVID-19(9). Here, we evaluated the association between COVID-19 outcomes and Vitamin D levels in patients attending a COVID-19 reference center in Mexico City. We aimed to identify determinants of Vitamin D levels in COVID-19 patients and develop causal-mediation models to propose mechanisms by which Vitamin D may lead to increased COVID-19 mortality.

## METHODS

### Study population

This study included consecutive patients evaluated at the Instituto Nacional de Ciencias Médicas y Nutrición Salvador Zubirán (INCMNSZ), a COVID-19 reference center in Mexico City between 17th March and 31st May 2020 with complete vitamin D measurements at admission(10). Subjects were initially assessed at triage and required either ambulatory or in-hospital care for COVID-19 (confirmed with computerized tomography (CT) and/or via RT-qPCR test from nasopharyngeal swabs. At the time of this writing, the INCMNSZ was a reference center for COVID-19 patients, which attended primarily severe and critical cases of COVID-19 from Mexico City. All patients had moderate to severe disease as defined by National Institute of Health criteria (*Moderate Illness:* Evidence of lower respiratory disease during clinical assessment or imaging and who have saturation of oxygen (SpO_2_) ≥94% on air. *Severe Illness:* SpO_2_ <94% on air, a ratio of arterial partial pressure of oxygen to fraction of inspired oxygen (PaO_2_/FiO_2_) <300 mm Hg, respiratory frequency >30 breaths/min, or lung infiltrates >50%). Subjects underwent a chest CT, and a radiologist determined the degree of pulmonary parenchymal disease and assessed epicardial fat thickness as a proxy for visceral fat. In addition, a medical history, anthropometric measurements, and laboratory tests were obtained, including 25 hydroxy-vitamin D. The electronic files of each patient were reviewed to document the outcomes during hospitalization. All proceedings were approved by the research and ethics committee of the INCMNZ (Ref 3383) and informed consent was waived due to the nature of the study.

### Laboratory and clinical measurements

Clinical variables and laboratory measures were obtained at the time of initial evaluation. Physical examination included: weight, height, body mass index (BMI, calculated as weight in kilograms divided by squared height in meters), pulse oximeter saturation (SpO2), respiratory rate (RR), temperature and arterial blood pressure (BP). Laboratory measurements included: full blood count and chemistry panel including liver function tests, C-reactive protein (CRP), fibrinogen, D-dimer, ferritin, troponin I (TPNI), erythrocyte sedimentation rate (ESR) and procalcitonin levels. The blood samples were processed in the central laboratory of the Instituto Nacional de Ciencias Médicas y Nutrición Salvador Zubirán, Vitamin D (25-hydroxivitamin D) was measured by chemiluminescence using the Abbott Architect I2000 equipment. Low levels of vitamin D were defined as <20ng/mL (<50 nmol/L) and deficient as a levels <12ng/mL (<30nmol/L) (6-9).

### Epicardial fat measurements

All patients underwent unenhanced CT scans including low-dose CT and two ultra-low-dose CT protocols with commonly reported imaging features of COVID-19 pneumonia. The thoracic CT was performed using a 64-slice scanner (GE MEDICAL SYSTEMS Revolution EVO). Epicardial adipose tissue (EAT) thickness was measured at 3 points (right atrioventricular fossa, left atrioventricular fossa, and anterior interventricular fossa) in the reformatted 4-chamber view using the multiplanar reconstruction (MPR) tool on the workstation(23–26). The maximum thickness of the EAT was determined from the surface of the myocardium to the pericardium (measured perpendicular to the pericardium). The measurements were made on 2 different occasions, obtaining a total of 6 measurements; the average of them was used for all statistical analyses. Pericardial adipose tissue (PAT) was quantified with the volume measurement tool with the Carestream system of the workstation. The thickness of the thoracic subcutaneous adipose tissue (TscAT) was measured from the anterior border of the sternum to the skin, at the level of the mitral valve in the axial plane of the tomography. The 80th gender-specific percentile of EAT thickness was obtained and used as the threshold to define increased EAT thickness. In addition, chest CT findings were recorded and used to evaluate severity of COVID infection(25).

### COVID-19 outcomes

Outcomes included mortality and critical disease (defined as the combination of mortality and need for mechanical ventilation /intubation). For time-to-event analyses, time from self-reported symptom onset prior to evaluation until last follow-up (censoring) or death, whichever occurred first was estimated.

### Statistical analyses

Cases with low and deficient vitamin D levels were analyzed using Student’s t-test or Mann-Whitney U according to the variable distribution (parametric or non-parametric) for continuous variables. The chi-squared test was applied for categorical variables. Logarithmic, squared root and cubic root transformations were carried out to ensure variable symmetry prior to modeling. Missing data on predictors other than Vitamin D were multiply imputed using the *mice* R package under the assumption of data missing completely at random and combined using Rubin’s rules. All statistical analyses were conducted using R software version 4.0.2.

### Predictors of vitamin D levels

Linear regression analyses were fitted to identify predictors of log-transformed vitamin D levels in patients with COVID-19, and model selection was carried out using minimization of the Bayesian Information Criterion (BIC). Logistic regression models were also fitted using a dummy variable which defined low and deficient vitamin D to identify predictors for these categories and again model selection was conducted using BIC. Finally, model diagnostics for linear regression were conducted using residual analyses and the Hosmer-Lemeshow test for logistic regression analyses.

### Prediction of mortality and severe COVID-19

Cox proportional risk regression analyses was used to investigate the association of vitamin D with mortality related to COVID-19. Univariate models and fully adjusted models were generated which included the following covariates: age, gender, BMI, C-reactive protein, D-dimer, ultrasensitive cardiac troponin, epicardial fat, T2D, CKD and oxygen saturation levels. An interaction effect was explored with BMI or BMI categories to rule out the differential impact of vitamin D adjusted for BMI. Model diagnostics were done using Schoenfeld residuals. Results are presented with Hazard Ratios (HR) and its corresponding 95% confidence intervals. Finally, the association of vitamin D with requirement for mechanical ventilation or the composite of critical COVID-19 was explored using logistic regression analyses adjusted for the covariates. Results are presented with Odds Ratios (OR) and its corresponding 95% confidence intervals.

### Causal mediation models

Finally, to explore whether variables which are influenced by Vitamin D levels may act as a mediator of the risk conferred by Vitamin D on COVID-19 severity, model-based causal mediation analyses were developed: D-dimer and ultrasensitive cardiac troponins were proposed as mediators of the effect of Vitamin D on COVID-19 mortality. All mediation analyses were performed using the mediation R package; to permit inference to obtain a 95% confidence interval using bias-corrected accelerated non-parametric bootstrap. To demonstrate the sequential ignorability assumption, a sensitivity analysis was run to demonstrate residual confounding by varying the correlation between the residuals of both the outcome and the moderator models. Statistical analyses were performed using R software version 4.0.3. A p value <0.05 was considered statistically significant

## RESULTS

### Study population

This study included 551 patients with confirmed COVID-19 (with compatible computerized tomography findings and/or positive RT-qPCR test from nasopharyngeal swabs) and vitamin D measurements. The mean age of participants was 51.92±13.74 years, with a male predominance (n=355, 64.4%), and a mean BMI of 30.05±5.72. Median follow-up was 15.0 days (IQR 10.0, 20.0) and 445 patients required hospitalization (81.1%). Overall, 93 patients received invasive mechanical ventilation (16.88%) and 116 in-hospital deaths (21.1%) were recorded. Type 2 diabetes (T2D) was present in 146 patients (26.9%), 219 patients had obesity (42.7%) and 217 were overweight (42.4%). Mean vitamin D levels were 21.78±9.01 and vitamin D levels below 20ng/mL were present in 251 subjects (45.6%) (**Table 1**). Extremely low vitamin D levels (≤12ng/mL) were observed in 59 patients (10.7%) (**Table 1 supplementary material**).

**Table 1.**
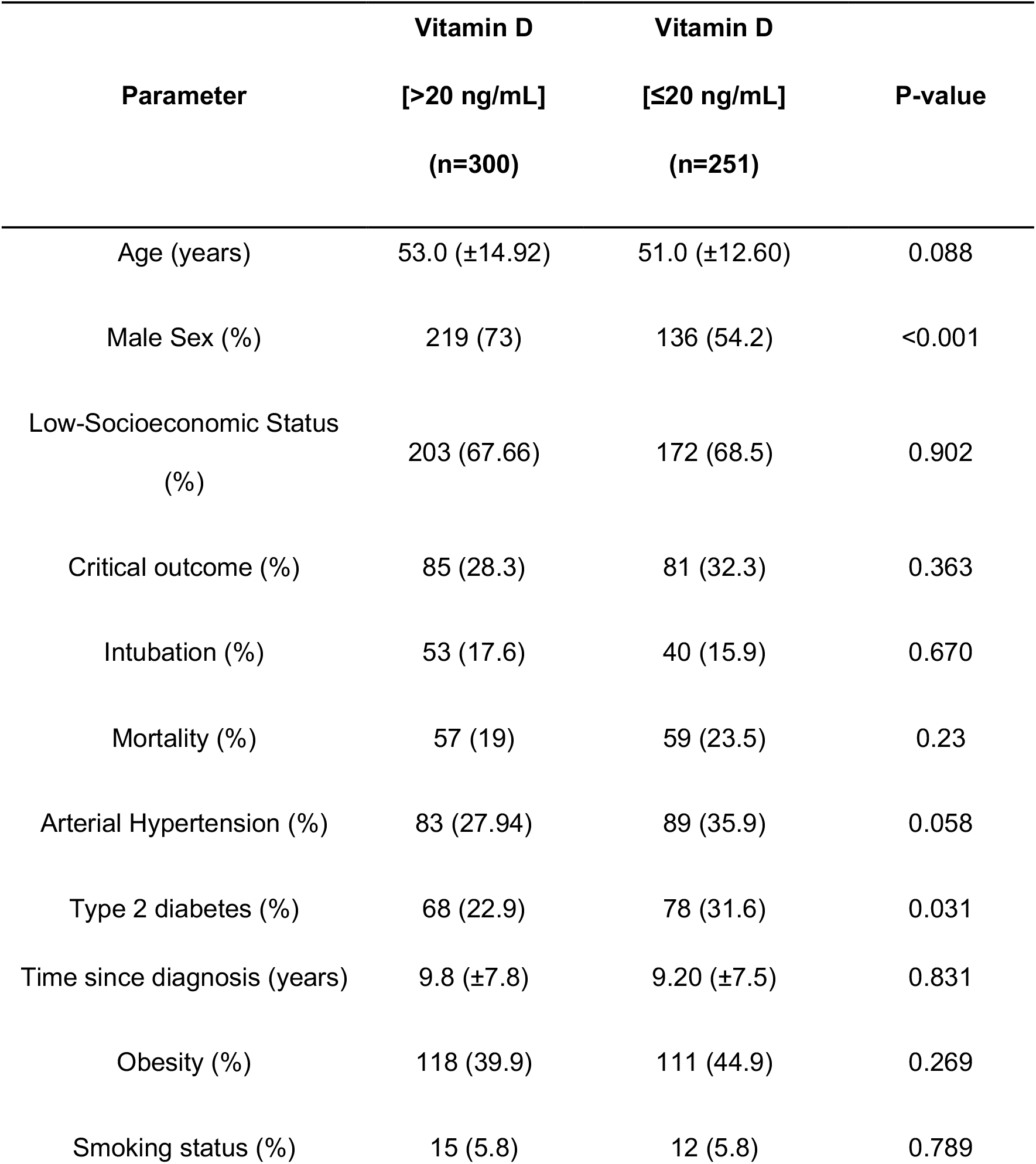
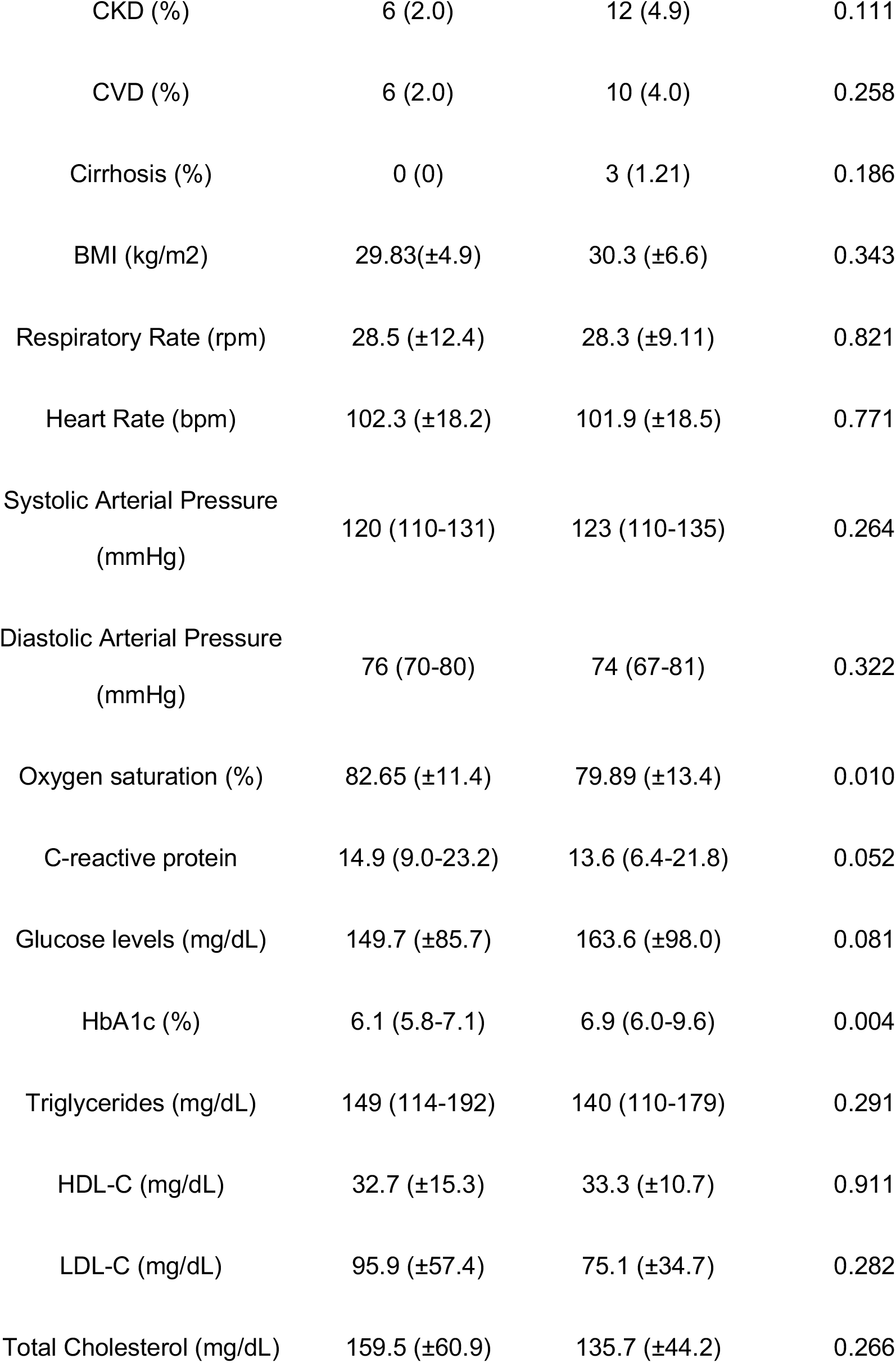
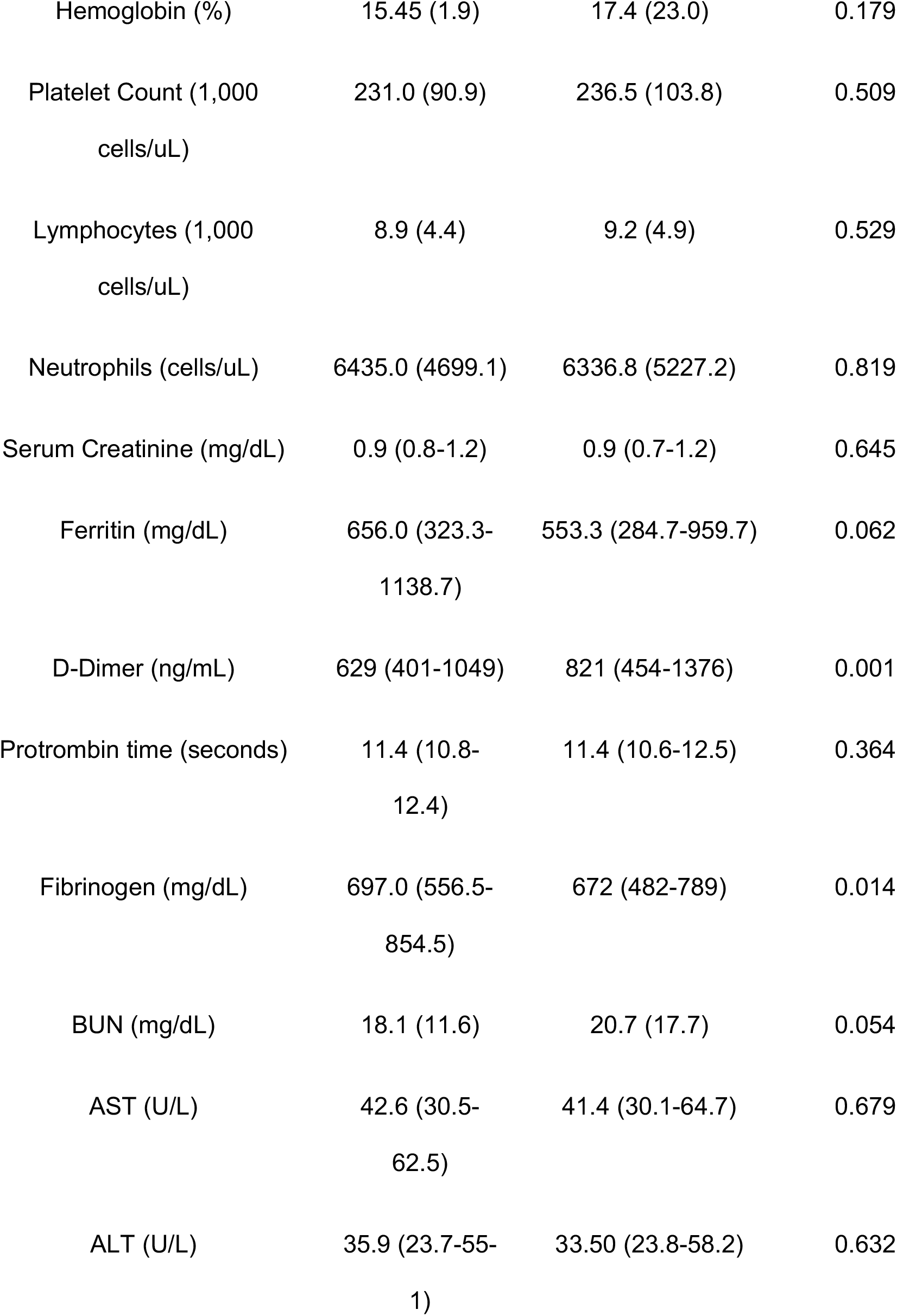
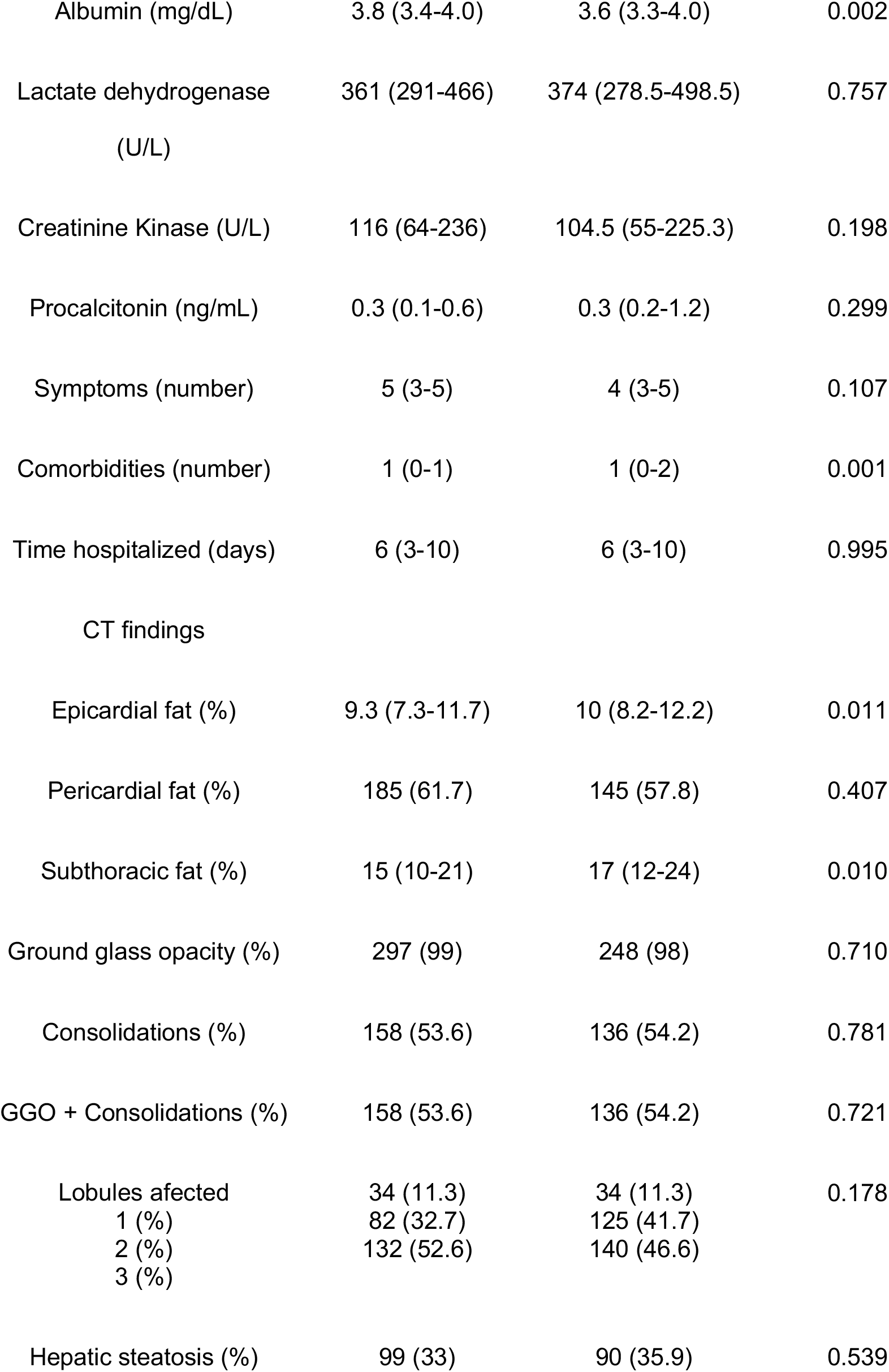
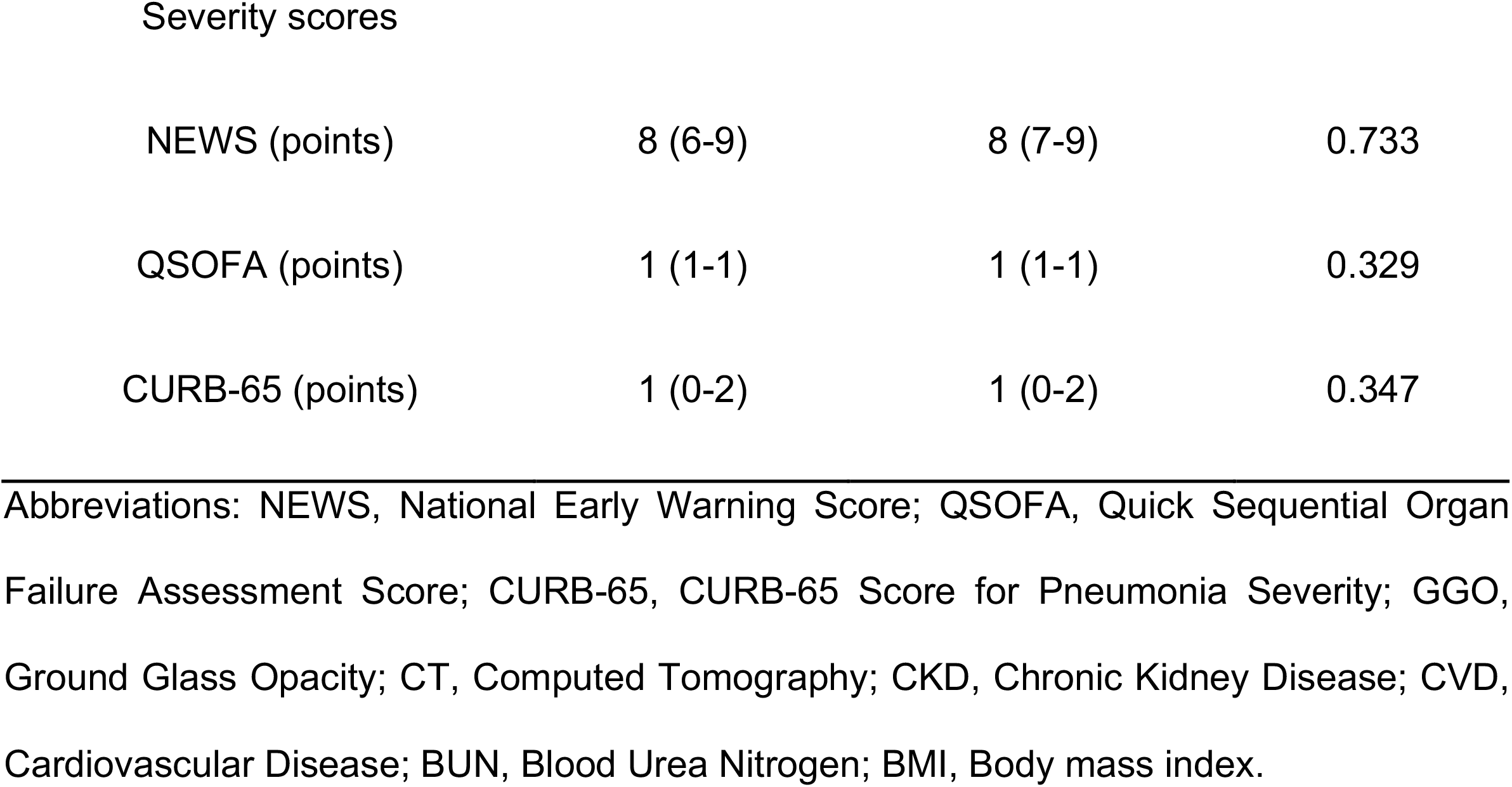
Clinical characteristics, imaging findings and severity scores in patients with COVID-19, comparing cases with and without low vitamin D levels. Values are presented as mean (± standard deviation) or median (inter-quartile range), where appropriate.

| Parameter | Vitamin D | Vitamin D | P-value |
| --- | --- | --- | --- |
| | [>20 ng/mL]<br>(n=300) | [≤20 ng/mL]<br>(n=251) | |
| Age (years) | 53.0 (±14.92) | 51.0 (±12.60) | 0.088 |
| Male Sex (%) | 219 (73) | 136 (54.2) | <0.001 |
| Low-Socioeconomic Status<br>(%) | 203 (67.66) | 172 (68.5) | 0.902 |
| Critical outcome (%) | 85 (28.3) | 81 (32.3) | 0.363 |
| Intubation (%) | 53 (17.6) | 40 (15.9) | 0.670 |
| Mortality (%) | 57 (19) | 59 (23.5) | 0.23 |
| Arterial Hypertension (%) | 83 (27.94) | 89 (35.9) | 0.058 |
| Type 2 diabetes (%) | 68 (22.9) | 78 (31.6) | 0.031 |
| Time since diagnosis (years) | 9.8 (±7.8) | 9.20 (±7.5) | 0.831 |
| Obesity (%) | 118 (39.9) | 111 (44.9) | 0.269 |
| Smoking status (%) | 15 (5.8) | 12 (5.8) | 0.789 |
| CKD (%) | 6 (2.0) | 12 (4.9) | 0.111 |
| CVD (%) | 6 (2.0) | 10 (4.0) | 0.258 |
| Cirrhosis (%) | 0 (0) | 3 (1.21) | 0.186 |
| BMI (kg/m<sup>2</sup>) | 29.83(±4.9) | 30.3 (±6.6) | 0.343 |
| Respiratory Rate (rpm) | 28.5 (±12.4) | 28.3 (±9.11) | 0.821 |
| Heart Rate (bpm) | 102.3 (±18.2) | 101.9 (±18.5) | 0.771 |
| Systolic Arterial Pressure<br>(mmHg) | 120 (110-131) | 123 (110-135) | 0.264 |
| Diastolic Arterial Pressure<br>(mmHg) | 76 (70-80) | 74 (67-81) | 0.322 |
| Oxygen saturation (%) | 82.65 (±11.4) | 79.89 (±13.4) | 0.010 |
| C-reactive protein | 14.9 (9.0-23.2) | 13.6 (6.4-21.8) | 0.052 |
| Glucose levels (mg/dL) | 149.7 (±85.7) | 163.6 (±98.0) | 0.081 |
| HbA1c (%) | 6.1 (5.8-7.1) | 6.9 (6.0-9.6) | 0.004 |
| Triglycerides (mg/dL) | 149 (114-192) | 140 (110-179) | 0.291 |
| HDL-C (mg/dL) | 32.7 (±15.3) | 33.3 (±10.7) | 0.911 |
| LDL-C (mg/dL) | 95.9 (±57.4) | 75.1 (±34.7) | 0.282 |
| Total Cholesterol (mg/dL) | 159.5 (±60.9) | 135.7 (±44.2) | 0.266 |
| Hemoglobin (%) | 15.45 (1.9) | 17.4 (23.0) | 0.179 |
| Platelet Count (1,000<br>cells/uL) | 231.0 (90.9) | 236.5 (103.8) | 0.509 |
| Lymphocytes (1,000<br>cells/uL) | 8.9 (4.4) | 9.2 (4.9) | 0.529 |
| Neutrophils (cells/uL) | 6435.0 (4699.1) | 6336.8 (5227.2) | 0.819 |
| Serum Creatinine (mg/dL) | 0.9 (0.8-1.2) | 0.9 (0.7-1.2) | 0.645 |
| Ferritin (mg/dL) | 656.0 (323.3-1138.7) | 553.3 (284.7-959.7) | 0.062 |
| D-Dimer (ng/mL) | 629 (401-1049) | 821 (454-1376) | 0.001 |
| Protrombin time (seconds) | 11.4 (10.8-12.4) | 11.4 (10.6-12.5) | 0.364 |
| Fibrinogen (mg/dL) | 697.0 (556.5-854.5) | 672 (482-789) | 0.014 |
| BUN (mg/dL) | 18.1 (11.6) | 20.7 (17.7) | 0.054 |
| AST (U/L) | 42.6 (30.5-62.5) | 41.4 (30.1-64.7) | 0.679 |
| ALT (U/L) | 35.9 (23.7-55-1) | 33.50 (23.8-58.2) | 0.632 |
| Albumin (mg/dL) | 3.8 (3.4-4.0) | 3.6 (3.3-4.0) | 0.002 |
| Lactate dehydrogenase<br>(U/L) | 361 (291-466) | 374 (278.5-498.5) | 0.757 |
| Creatinine Kinase (U/L) | 116 (64-236) | 104.5 (55-225.3) | 0.198 |
| Procalcitonin (ng/mL) | 0.3 (0.1-0.6) | 0.3 (0.2-1.2) | 0.299 |
| Symptoms (number) | 5 (3-5) | 4 (3-5) | 0.107 |
| Comorbidities (number) | 1 (0-1) | 1 (0-2) | 0.001 |
| Time hospitalized (days) | 6 (3-10) | 6 (3-10) | 0.995 |
| CT findings |  |  |  |
| Epicardial fat (%) | 9.3 (7.3-11.7) | 10 (8.2-12.2) | 0.011 |
| Pericardial fat (%) | 185 (61.7) | 145 (57.8) | 0.407 |
| Subthoracic fat (%) | 15 (10-21) | 17 (12-24) | 0.010 |
| Ground glass opacity (%) | 297 (99) | 248 (98) | 0.710 |
| Consolidations (%) | 158 (53.6) | 136 (54.2) | 0.781 |
| GGO + Consolidations (%) | 158 (53.6) | 136 (54.2) | 0.721 |
| Lobules affected | 34 (11.3) | 34 (11.3) | 0.178 |
| 1 (%) | 82 (32.7) | 125 (41.7) |  |
| 2 (%) | 132 (52.6) | 140 (46.6) |  |
| 3 (%) |  |  |  |
| Hepatic steatosis (%) | 99 (33) | 90 (35.9) | 0.539 |
| Severity scores |  |  |  |
| NEWS (points) | 8 (6-9) | 8 (7-9) | 0.733 |
| QSOFA (points) | 1 (1-1) | 1 (1-1) | 0.329 |
| CURB-65 (points) | 1 (0-2) | 1 (0-2) | 0.347 |

Abbreviations: NEWS, National Early Warning Score; QSOFA, Quick Sequential Organ Failure Assessment Score; CURB-65, CURB-65 Score for Pneumonia Severity; GGO, Ground Glass Opacity; CT, Computed Tomography; CKD, Chronic Kidney Disease; CVD, Cardiovascular Disease; BUN, Blood Urea Nitrogen; BMI, Body mass index.

### Determinants of Vitamin D levels amongst patients with COVID-19

The pathophysiological adaptations to COVID-19 may be predictive of low vitamin D levels. To this end, determinants of low vitamin D in COVID-19 were sought in an attempt to develop a mechanistic explanatory model for this relationship. Subjects with low vitamin D levels (<20mmg/dl) were more likely to be female, have type 2 diabetes, higher HbA1c, D-dimer and ferritin levels and lower oxygen saturation, albumin and C-reactive protein. Using linear regression, a lower log-transformed vitamin D level was independently associated with female gender, higher log-transformed ultrasensitive cardiac troponin, higher log-transformed D-dimer, higher log-transformed epicardial fat area, and lower C-reactive protein levels (**Table 2**). When exploring a model to detect low vitamin D levels, there was a significantly higher odds for log-transformed D-dimer levels (OR 1.31, 95%CI 1.06, 1.63); lower odds were associated with male gender (OR 0.45, 95%CI 0.31, 0.65), higher oxygen saturation levels (OR 0.98, 95%CI 0.97, 0.99) and higher C-reactive protein values (OR 0.75, 95%CI 0.61, 0.92), adjusted for age, and log-transformed epicardial fat. There was no association between days from symptom onset and vitamin D levels at admission.

**Table 2.**
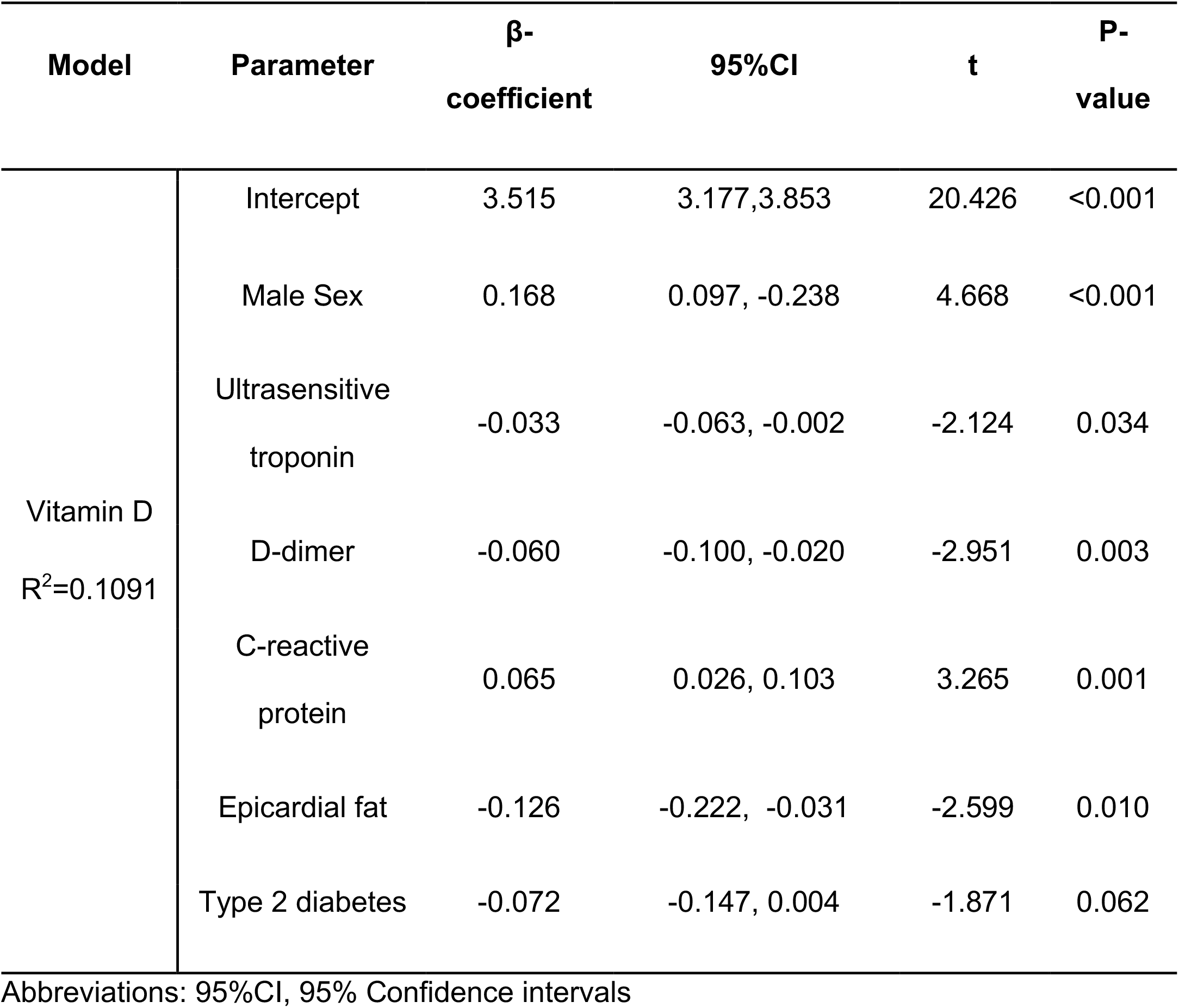
Multiple linear regression model to identify determinants of Vitamin D levels in patients with COVID-19.

### Vitamin D levels and COVID-19 mortality

Overall, vitamin D levels were significantly lower when comparing non-fatal to fatal COVID-19 cases (22.41±9.34 vs. 19.44±67.19, p<0.001). When assessing risk related to the association between mortality and vitamin D levels using Cox regression, a 1-unit increase in vitamin D levels was associated with a decreased risk of COVID-19 mortality. Interestingly, when stratifying cases according to gender, the difference in vitamin D levels between fatal and non-fatal cases was greater in women compared to men and lower in cases with obesity (**Figure 1**). When the mortality models were adjusted for age, gender, BMI, and C-reactive protein, CKD and T2D the observed association between vitamin D levels and a decrease in COVID-19 mortality persisted (**Table 3**). There was no significant interaction with BMI, (as a continuous variable or categorized) in normal weight, overweight and obese with vitamin D levels. Using post-estimation simulation to predict risk associated with changes in vitamin D levels using the simPH R package, there was a steady decrease in risk attributable to increasing vitamin D concentrations using vitamin D <20ng/mL and ≤12ng/mL as thresholds (**Figure 2**).

**Table 3.** Cox proportional risk regression models to predict mortality related to COVID-19 using Vitamin D levels adjusted for covariates. Model 1: Unadjusted, Model 2: Adjusted for age, gender, body-mass index (BMI) and C-reactive protein (CRP), Model 3: Model 2 adjusted for chronic kidney disease (CKD), epicardial fat and type 2 diabetes (T2D).

| Model | Parameter | β-<br>coefficient | HR (95%CI) | P-value |
| --- | --- | --- | --- | --- |
| Model 1<br>c-statistic=0.566 | Vitamin D | -0.036 | 0.965 (0.942, 0.988) | 0.003 |
| Model 2<br>c-statistic=0.694 | Vitamin D | -0.043 | 0.938 (0.934, 0.983) | 0.001 |
|  | Age | 0.037 | 1.037 (1.022, 1.052) | <0.001 |
|  | Male gender | 0.787 | 2.196 (1.373, 3.512) | 0.001 |
|  | BMI | 0.038 | 1.039 (1.005, 1.073) | 0.024 |
|  | CRP | 0.004 | 1.004 (1.002, 1.007) | <0.001 |
| Model 3<br>c-statistic=0.702 | Vitamin D | -0.039 | 0.962 (0.935, 0.989) | 0.006 |
|  | CKD | 0.301 | 0.749 (0.276, 2.030) | 0.570 |
|  | T2D | 0.325 | 1.384 (0.882, 2.170) | 0.157 |
|  | Ultrasensitive<br>cardiac<br>troponin | 0.170 | 1.185 (1.002, 1.403) | 0.048 |
|  | D-dimer | 0.003 | 1.003 (0.804, 1.252) | 0.980 |
|  | Oxygen saturation | -0.026 | 0.975 (0.960, 0.990) | 0.001 |
|  | Epicardial fat thickness | 0.669 | 1.952 (0.501, 7.607) | 0.335 |

Abbreviations: CKD, Chronic Kidney Disease; BMI, Body mass index; CRP, C-reactive protein; T2D, Type 2 diabetes mellitus; HR, Hazard Ratio; 95%CI, 95% Confidence Interval

**Table 4:** Causal mediation analyses predicting the effect of Vitamin D levels (E) mediated by elevated D-dimer, ultrasensitive cardiac troponins or low SpO2 (M) on severe COVID-19 and mortality (Y), adjusted by gender, age, BMI and epicardial fat. **<u>Abbreviations</u>**: ACME: average causal mediation effects; ADE: average direct effects; SpO2: Oxygen saturation levels.

| Outcome (Y) | Efactor (E) | Mediator<br>(M) | ACME<br>(95%CI) | ADE<br>(95%CI) | Total Effect<br>(95%CI) | Proportion<br>Mediated<br>(95%CI) |
| --- | --- | --- | --- | --- | --- | --- |
| COVID-19<br>Mortality | Vitamin D | D-dimer | -0.035 | -0.144 | -0.179 | 19.3% |
|  |  |  | (-0.069,-0.010) | (-0.164, -0.10) | (-0.188,-0.050) | (9.5-77.0%) |
|  |  | Ultrasentivie<br>cardiac troponins | -0.047 | -0.133 | -0.181 | 26.2% |
|  |  |  | (-0.085, -0.020) | (-0.150, -0.020) | (-0.183, -0.060) | (15.0-73.0%) |
|  |  | SpO2 | -0.037 | -0.144 | -0.181 | 20.3% |
|  |  |  | (-0.066, -0.010) | (-0.159, -0.030) | (-0.186,-0.050) | (9.7-64.0%) |

**Figure 1.**
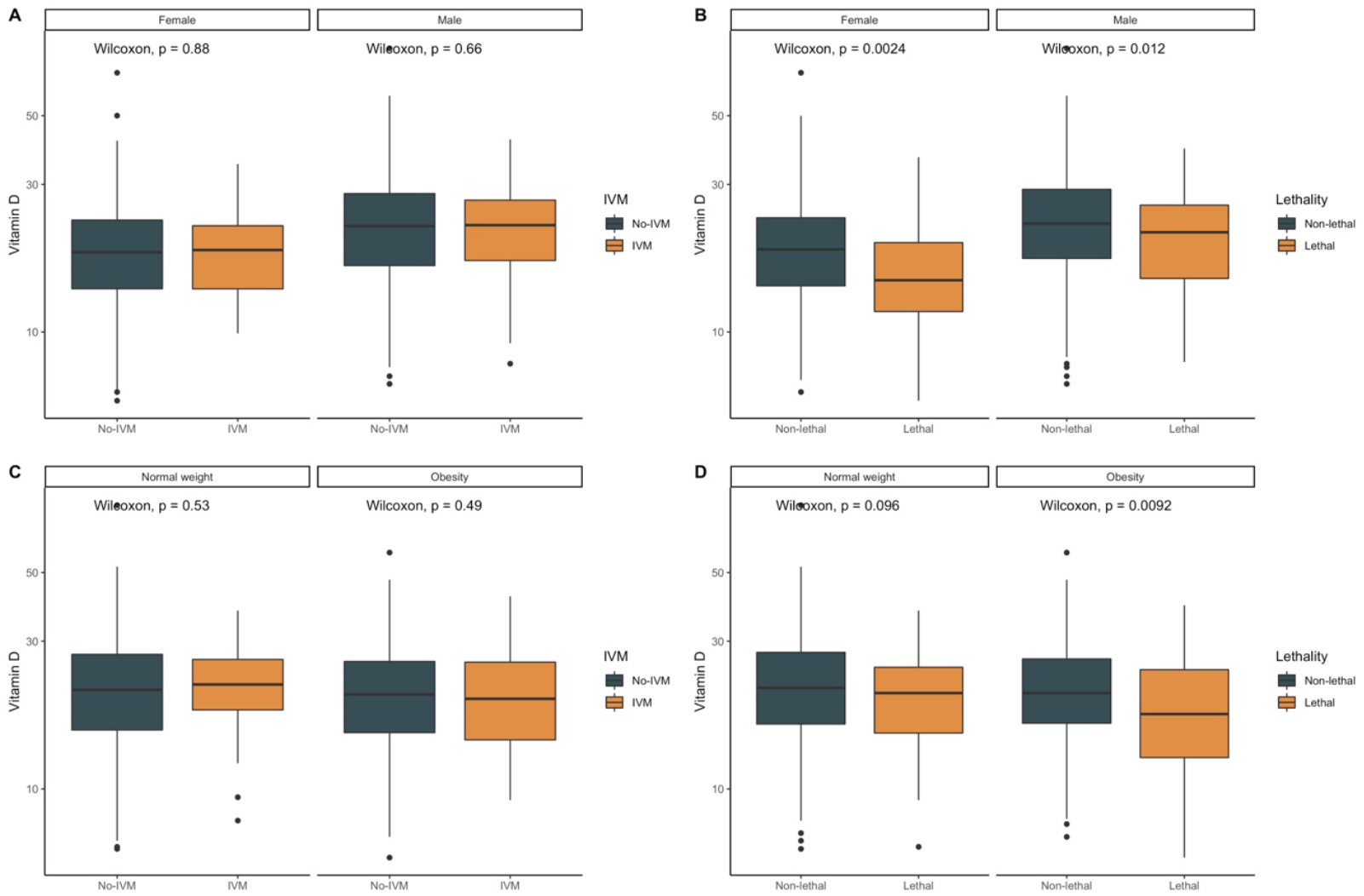
Boxplots comparing Vitamin D levels according to the need for invasive mechanical ventilation (IVM) or lethal COVID-19 stratified by gender (A-B) and by body-mass index categories (C-D).

**Figure 2.**
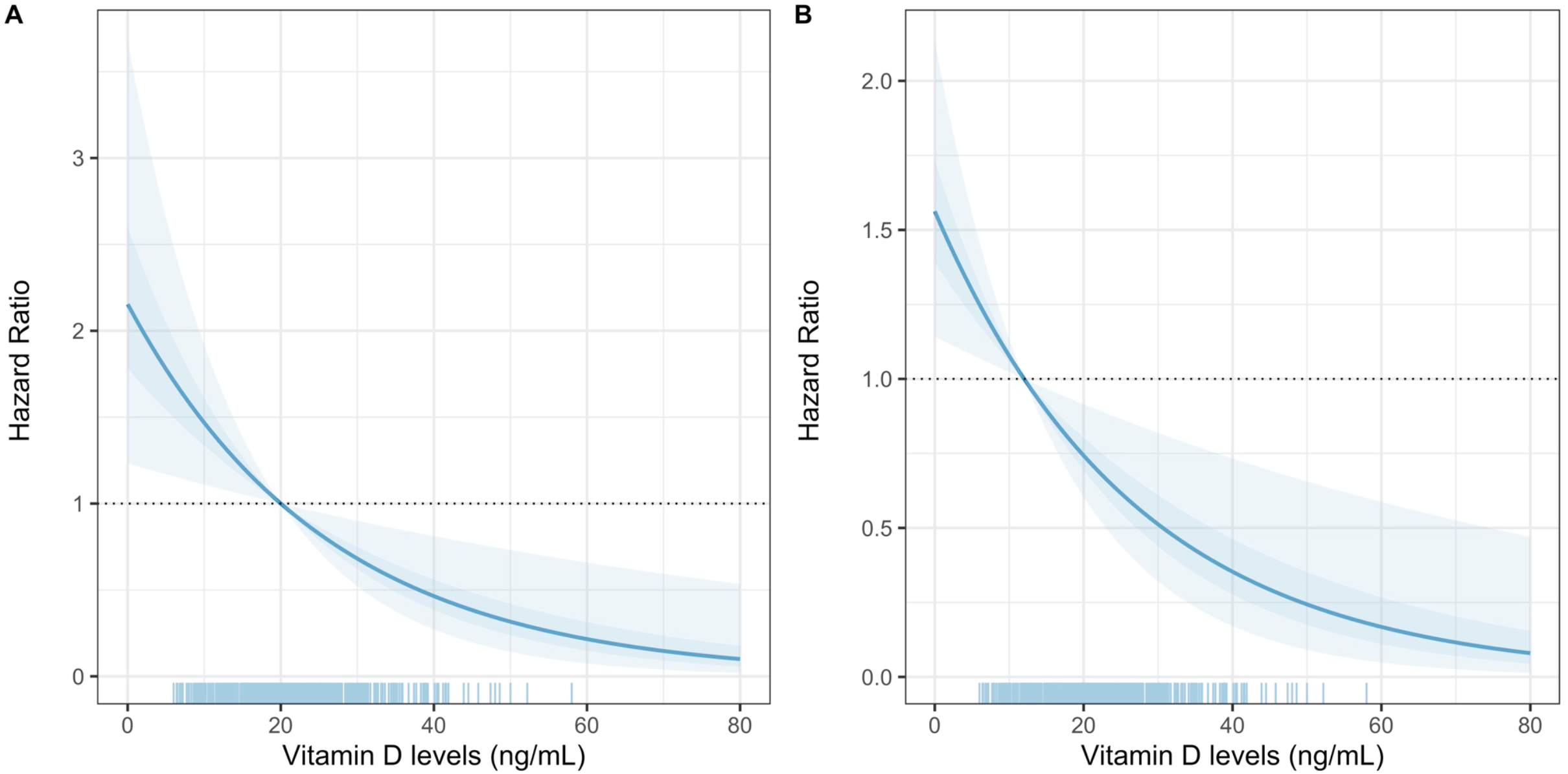
Post-estimation simulation of Vitamin D levels to predict COVID-19 lethality, adjusted for age, sex, BMI, C-reactive protein, epicardial fat, D-dimer, oxygen saturation, T2D, and CKD using Vitamin D cut-offs of <20ng/dL (A) and ≤12ng/mL

**Figure 3.**
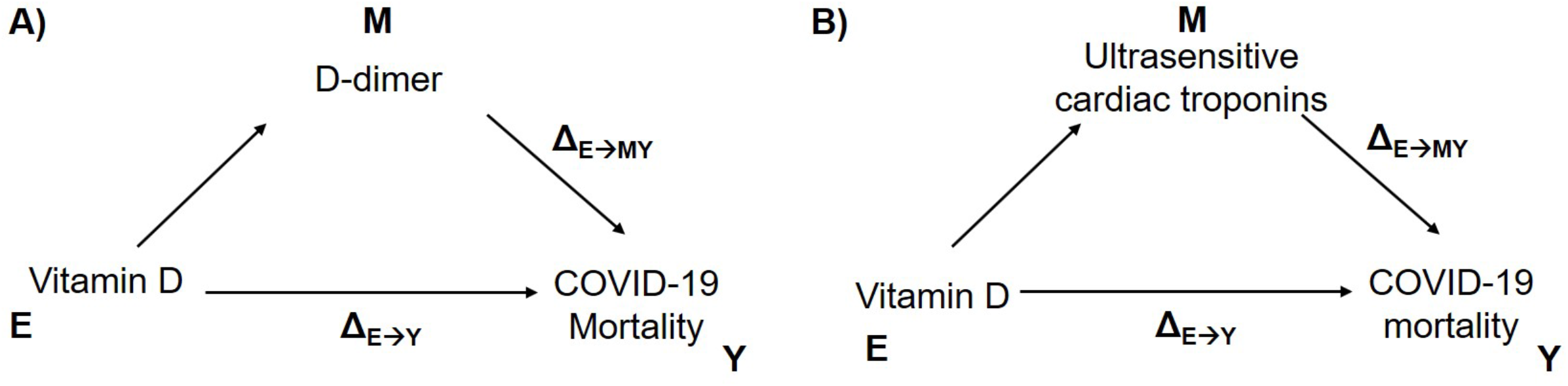
Model-based causal mediation analyses to investigate the role of Vitamin D on COVID-19 mortality mediated by D-dimer (A), ultrasensitive cardiac troponins (B)

### Vitamin D and critical COVID-19

When assessing the impact of vitamin D on the risk of invasive mechanical ventilation there was no significant association even after adjustment for age, gender, BMI, C-reactive protein, CKD or T2D status (OR 0.986, 95%CI 0.957, 1.015, p=0.366). However, when assessing the composite of critical COVID-19 using logistic regression models, lower vitamin D levels were associated with critical COVID-19 (OR 0.97, 95%CI 0.94, 0.99, p=0.042, adjusting by age, gender, BMI, C-reactive protein, D-dimer, CKD, SpO2 or T2D status).

### Causal mediation models

Finally, model-based causal mediation models were developed to assess whether the effect of vitamin D (E) on increased mortality risk (Y) was mediated through changes in variables identified in **Table 2** (M), adjusted for age, gender, BMI and epicardial fat. The direct effect of vitamin D on increased mortality risk was significant (Δ_E→Y_ -0.144, 95%CI -0.069, -0.010) and the indirect effect of vitamin D, mediated by increase D-dimer levels (Δ_E→MY_ -0.035, 95%CI -0.164, -0.010), represented 19.3% (95%CI 9.5, 77.0%) of the overall association of vitamin D on mortality. A similar scenario was observed for cardiac troponins, whereby both the direct effect of vitamin D on mortality (Δ_E→Y_ -0.133, 95%CI -0.150, -0.020) and the indirect effect mediated by ultrasensitive cardiac troponins (Δ_E→MY_ -0.047, 95%CI -0.085, - 0.020), were significant and represented 26.2% (95%CI 14.9, 73.0%) of the overall effect of vitamin D on mortality. Notably, there were no significant causal mediation models for either BMI or epicardial fat, here there was only a direct effect on mortality, independent of vitamin D levels.

## DISCUSSION

In this study, the association between vitamin D levels and severity of COVID-19 was explored in a Mexican population. Deficient levels of vitamin D (deficiency) showed a clear association with mortality, even after adjusting for confounders, including epicardial fat as a proxy of visceral fat and BMI. A vitamin D level <20ng/mL (<50nmol/L), showed a strong negative predictive value, suggesting that when levels are adequate, the probability of mortality is low. Furthermore, the increased risk of mortality from COVID-19 was partly mediated by the effect of vitamin D on markers of disease severity, such as D-dimer and ultrasensitive cardiac troponins, independent of BMI and epicardial fat (these showed effects on COVID-19 mortality independent of vitamin D levels). This suggests that vitamin D may be a marker of an impaired response to infection within the pulmonary epithelium, most notably in those with severe deficiency(27).

Several studies have explored the relationship between COVID-19 and vitamin D levels(18,20,27,28). These include those examining vitamin D levels and risk of infection and those examining an association with severity of COVID-19. Higher levels of IL-6 were observed in vitamin D deficiency suggesting a greater inflammatory response in these patients(29,30). A recent systematic review and meta-analysis reported that vitamin D deficiency was not associated with increased risk of infection, but severe cases presented with greater vitamin D deficiency compared with mild cases. Vitamin D deficiency has been associated to increased hospitalization and mortality risk from COVID-19(16,18). Physiological mechanisms by which vitamin D exerts a protective function include enhanced innate immunity including augmentation of physical barriers to infection and optimization of adaptive immunity(27). Vitamin D has also been proposed to exert a modulatory effect on the inflammatory response caused by COVID-19 by curbing adaptive immunity though inhibition og B cell proliferation, differentiation and production of antibodies and plays a role in regulation the T cell phenotype. Thus, there is a shift in the adaptive immune response from Th1 to a more regulatory Th2 response, characterized by an increase in expression of Th2 associated cytokines. This may attenuate the quantity of pro-inflammatory cytokines that are associated with severe infection(27). In addition, vitamin D induces ACE-2 expression, and suppresses the angiotensin-renin system, thus reducing levels of proinflammatory angiotensin II. Hence, vitamin D deficiency, could potentiate the cytokine storm perpetuating a pro-inflammatory state and worsening pulmonary outcomes(28,29). Finally, thrombotic complications are common in such patients; vitamin D is also involved in the regulation of thrombotic pathways(31,32). The mechanisms through which Vitamin D may influence pro-thrombotic pathways is closely related to its anti-inflammatory properties, which reduce endothelial activation and oxidative stress(33). In our study, low vitamin D levels in COVID-19 patients were related to inflammatory, pro-thrombotic and metabolic markers of severity, confirming observations from previous studies.

Adverse COVID-19 outcomes have been linked to the ethnic origin of the population under study and its socio-economic characteristics; this has also been associated with the presence of vitamin D deficiency. Asians, African Americans, and ethnic minorities are at an increased risk of mortality from COVID-19(34). This may partly be due to a decrease in the production of vitamin D dependent on UV rays. This is related to the skin levels of melanin present in these populations and on the unequal distribution of poverty and cardiometabolic disease rates across such ethnicities and populations. This finding is relevant and may explain lower vitamin D levels and severity of COVID-19 in México in previous studies(35–37). Previous reports in similar populations, including Hispanics, have shown higher risk of severe SARS-CoV-2 infection, compared to Caucasian population(38). This could be attributable to increased cardio-metabolic comorbidities in cases from Mexico, where vitamin D deficiency is more prevalent in type 2 diabetes and obesity primarily due to increased adiposity, as supported by our results(39). The relationship between vitamin D levels and sex may also underlie our finding of lower vitamin D levels in women, who have increased adiposity content compared to males(40). Low vitamin D levels may also occur in CKD, by reduced expression of 1-alpha hydroxylase; given a large, despite its influce on Vitamin D status, our study only included a reduced number of cases with CKD. Currently, routine vitamin D supplementation in hospitalized patients with COVID-19 is not recommended. A recent clinical trial study study showed that administration of a high dose of calcifediol or 25-hydroxyvitamin D, did not reduce length of hospital stay in patients COVID-19 (20). Ideally, additional large randomized controlled trials are needed to properly assess this claim and whether vitamin D supplementation can significantly impact risk of severe COVID-19.

This study has certain strengths and limitations. It included a large sample of patients with heterogenous risk profiles in whom a variety of disease severity parameters were measured. In addition, a series of statistical tests were carried out to ensure minimal possibility of residual confounding. Nevertheless, some limitations must be acknowledged to properly interpret this study. First, a chemiluminescence immunoassay was used to assess vitamin D levels, this may lead to inconsistent results compared to other techniques including competitive binding protein - CBP, radioimmunoassay - RIA liquid chromatography - LC, UV detection with liquid chromatography and liquid chromatography mass spectrometry LCMS or tandem mass spectrometry. Since most patients were attended in the institution for the first time for COVID-19, historic vitamin D values were not available to assess the effect of vitamin D dynamics on infection risk or outcomes. Furthermore, given the disease course of COVID-19, severity profiles are highly heterogeneous even amongst hospitalized patients, which may influence vitamin D values based on varying severity; control for this factor using propensity score matching was carried out, however, there remains a possibility for residual confounding. Finally, since this is a secondary analysis, post-hoc sample size calculation was not performed, and negative results should be interpreted with caution. Notably, these results are from a COVID-19 reference center in Mexico City, this could reduce the representativeness of the findings primarily to severe and critical forms of COVID-19 from the central region of Mexico. Further evidence in other regions of Mexico to confirm the role of vitamin D as a marker of disease severity and mortality in Mexicans with COVID-19 is needed.

### Conclusions

Vitamin D levels ≤12 ng/ml (30nmol/L) are independently associated with COVID-19 mortality, even after adjusting for confounders, including measures of visceral and total body fat. No association was confirmed between vitamin D levels and the need for intubation. Vitamin D deficiency is more prevalent in women and patients with type 2 diabetes mellitus. Vitamin D supplementation may be considered in deficient patients, but evidence of benefit is required from double blind randomized controlled trials.

## Data Availability

Data is available from the corresponding author upon reasonable request. Code for reproducibility of results available at: https://github.com/oyaxbell/covid_metabolism

## ACKNOWLEDGMENTS

All authors approved the submitted version. All the authors would like to thank the staff of the Endocrinology and Metabolism Department for all their support. We are thankful to the study volunteers for all their work and support throughout the realization of the study. AVV, and NEAV are enrolled in the PECEM program at the Faculty of Medicine of UNAM; AVV and NEAV are supported by CONACyT.

## CONTRIBUTIONS

research idea and study design: RM, OYBC, CAAS; data acquisition: RM, PEVC, NRP, MAJA, CIPC, BRE, JCVG, CAAS; data analysis/interpretation: OYBC, RM, CAAS, NEAV, AVV; statistical analysis: OYBC, NEAV; manuscript drafting: RM, OYBC, NEAV, AVV, CAAS; supervision or mentorship: RM, CAAS, APL, JSO. Each author contributed important intellectual content during manuscript drafting or revision and accepts accountability for the overall work by ensuring that questions pertaining to the accuracy or integrity of any portion of the work are appropriately investigated and resolved.

